# Longitudinal analysis of antibody responses to the Pfizer BNT162b2 vaccine in Patients Undergoing Maintenance Hemodialysis

**DOI:** 10.1101/2021.07.20.21260849

**Authors:** André Weigert, Marie-Louise Bergman, Lígia Gonçalves, Iolanda Godinho, Nádia Duarte, Rita Abrantes, Patrícia Borges, Ana Brennand, Vanessa Malheiro, Paula Matoso, Onome Akpogheneta, Lindsay Kosack, Pedro Cruz, Estela Nogueira, Magda Pereira, Ana Ferreira, Marco Marques, Telmo Nunes, João Viana, Jocelyne Demengeot, Carlos Penha-Gonçalves

**Affiliations:** DaVita Óbidos, Casais de Alvito, Portugal; Serviço de Nefrologia, Hospital Santa Cruz, Centro Hospitalar de Lisboa Ocidental, Carnaxide, Portugal; Instituto de Farmacologia e Neurociências, Faculdade de Medicina, Universidade de Lisboa, Lisboa, Portugal; IGC, Instituto Gulbenkian de Ciência, Oeiras, Portugal; Serviço de Nefrologia e Transplantação Renal, Centro Hospitalar de Lisboa Norte EPE, Lisboa, Portugal; Serviço de Nefrologia, Centro Hospitalar do Médio Tejo, Torres Novas, Portugal; Serviço de Nefrologia, Hospital das Forças Armadas, Lisboa, Portugal; Affidea Laboratório Lisboa, Portugal; CIISA, Centro de Investigação Interdisciplinar em Sanidade Animal, Faculdade de Medicina Veterinária, Universidade de Lisboa, Lisboa, Portugal; Serviço de Patologia Clínica, Centro Hospitalar de Lisboa Ocidental EPE, Carnaxide, Portugal

**Keywords:** BNT162b2, Chronic hemodialysis, COVID-19, IgG, SARS-CoV-2, Vaccine

## Abstract

**Background:** Hemodialyzed patients are at higher risk for COVID-19 and were prioritized in the Portuguese vaccination campaign

**Methods:** We performed a prospective, longitudinal, cohort analysis of 143 patients on hemodialysis and 143 age-matched controls along BTN162b2 vaccination. ELISA quantified anti-full-length Spike IgG, IgM and IgA levels prior to the first vaccine dose (t0); 3 weeks later (second dose, t1); and 3 weeks later (t2); 127 patients were re-evaluated140 (t3) and 180 days (t4) after the first dose.

**Results:** Seroconversion at t1 was remarkably low in patients, with positivity for anti-spike IgG, IgM and IgA antibodies of 29.4%, 12% and 41%, respectively, increasing to 90.9% (IgG) and 83.9% (IgA) in t2, (IgM remained unchanged). Below 70 years of age anti-spike IgG levels at t1 were significantly lower compared to age-matched controls and showed a profile similar to older individuals. Immunosuppression was associated with lower antibody responses (p=0.005 at t1; p=0.008 at t2). Previous unresponsiveness to hepatitis B vaccination (75/129, 58% of patients negative for anti-HBs antibodies) did not correlate with humoral unresponsiveness to BTN162b2. Anti-spike IgG, IgM and IgA positivity and antibody levels significantly decay at t3, with IgG levels showing further waning at t4.

**Conclusions:** The large majority of hemodialyzed patients showed IgG seroconversion upon BNT162b2 mRNA vaccination, albeit a sizable proportion of patients presented poor responses. Follow-up of antibody responses 180 days post vaccination unveiled significant decay of anti-spike antibodies and warrant close monitoring of COVID-19 infection and further studies on reinforced vaccination schedules in patients undergoing maintenance hemodialysis.

## INTRODUCTION

Patients receiving in-center hemodialysis treatment are at increased risk of SARS-Cov-2 infection. Mortality of hemodialyzed patients with COVID-19 is higher (1) and they may pose an additional stress in hospital’s dialysis capacity when admitted, as most of these patients receive routine dialysis treatments as outpatients. Although vaccination has been widely recommended to this specific population, the efficacy in long term antibody generation and the effectiveness in reducing transmission and severe infection is still uncertain. The reduction of 90% in the incidence of COVID-19 after vaccination with the Pfizer BNT162b2 vaccine in the general population (2,3) or in SARS-Cov-2 RNA detection in vaccinated versus non-vaccinated health workers (4) might not be reproduced in vaccinated hemodialysis patients. Initial studies revealed success in antibody generation but reduced titers in comparison with healthy controls (5–7). We evaluated the IgG, IgM and IgA anti-spike antibody response along the vaccination schedule defined for BTN162b2 mRNA vaccine and characterized responders and non-responders in our cohort of hemodialysis outpatients. Furthermore, medium term antibody response was performed 140 and 180 days after vaccination. In addition, responses to the BTN162b2 vaccine were compared to the hepatitis B vaccination response and to total immunoglobulin titers.

## MATERIAL AND METHODS

### ETHICS STATEMENT

This study is covered by approvals from the Ethics committees of DaVita in Portugal, Centro Hospitalar Lisboa Ocidental and the Administração Regional de Lisboa e Vale do Tejo in compliance with the Helsinki Declaration of 1975, as revised in 2013, and follows international and national guidelines for health data protection. All participants provided informed consent to take part in the study.

### STUDY DESIGN

The study enrolled 156 patients with stage 5 chronic kidney disease (CKD) undergoing renal replacement therapy as outpatients at a hemodialysis clinic (DaVita, Eurodial) in Óbidos, Portugal. An age-matched control cohort without kidney disease comprised 143 individuals randomly selected from a larger cohort of 1245 Health care workers and 146 nursing home residents (8). All Participants initiated BNT162b2 mRNA vaccination (Comirnaty®, Pfizer/BioNTech) according the established schedule of 2 doses with a 3 weeks interval. Venous blood was collected at the day of first vaccine dose (time 0, t0), 3 weeks later at the day of the second dose (t1), and 3 weeks after the second dose (t2) (**Figure 1A and B**). Participants with evidence of COVID-19 infection were excluded [serum reactivity against SARS-CoV-2 nucleocapsid (N) at time of enrolment (n=3) or SARS-CoV-2 RNA positivity in RT-PCR test before enrolment (n=2) or during the collection time (n=3)]. Between t0 and t1, two patients died and two patients dropped-out of the study. Between t1 and t2, one patient was hospitalized with non-COVID-19 respiratory infection (**Figure 1A**). To follow-up antibody responses venous blood was collected from 127 patients at 140 days (t3) and 180 days (t4) post first vaccine dose. Clinical data was collected from medical records and dedicated questionnaire.

**Figure 1.**
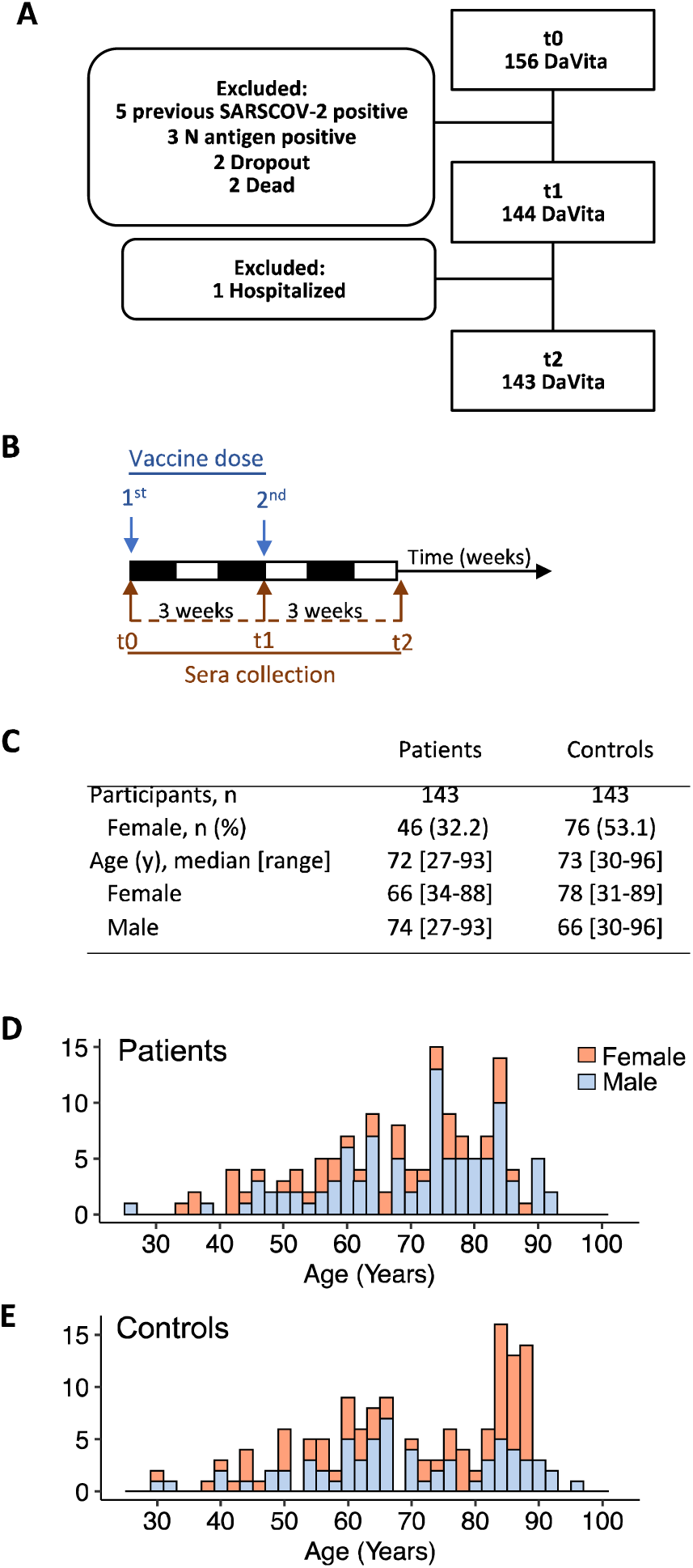
Óbidos hemodialysis and control cohort. A) Patient enrolment along the vaccination schedule, exclusion criteria and drop-outs. B) Serum collection schedule during vaccination with BNT162b2 RNA was performed at 3 time points: at the time of first dose inoculation (t0); 3-5 weeks after the first dose (t1) and; 3 weeks after the second dose (t2). C-E) Age and sex profiles of final sample in patients and controls.

### ANTIBODY MEASUREMENTS

The ELISA assay used to quantify IgG, IgM and IgA anti-full-length SARSCoV2 spike was adapted from (9) and semi-automized in a 384-well format, according to a protocol to be detailed elsewhere. Assay performance was determined by testing 1000 pre-pandemic sera and 40 COVID-19 patients diagnosed at least 10 days prior to sera collection. ROC curve analysis determined a specificity of 99.3%, 99.2%, 99.2%, and a sensitivity of 95.9%, 61.2%, 73.7% for IgG, IgM and IgA, respectively. Individual assay readouts (OD values) were standardized using as calibrators, samples obtained from COVID-19 confirmed patients, in all ELISA plates, and the normalized OD (ODnorm) adjusted to set ODnorm=1 as positivity cut-off. Serial titration of 67 COVID-19 patients established that the assay is semi-quantitative and has a dynamic range of 3 logs titer. Each sample was assayed in duplicates and any identified discrepancies resolved by repeating the test. Antibodies against SARS-Cov-2 N antigen were measured by an electrochemiluminescence immunoassay (ECLIA), from Roche Diagnostics (Elecsys^®^ Anti-SARS-CoV-2) and total IgG, IgM and IgA at t2 were quantified using three immunoturbidimetric methods (PEG enhanced) from Siemens Healthineers, using Siemens Atellica CH Analyzer, following manufacturer instructions.

### STATISTICAL ANALYSIS

Quade test was used to analyze individuals in temporal series and pairwise group comparisons between different time points used the Wilcoxon signed-rank test **(Figure 2C and Figure 3)**. Mann-Whitney U Test (Wilcoxon Rank Sum Test) was used for pairwise comparison between the age groups or single time point comparisons (**Figure 2C, Figure 3 and Figure 6**). To test for effects clinical conditions within a specific group on the magnitude of the antibody class responses, the Wilcoxon rank sum test was used (**Figure 4 and Supplemental Figure 1**). Fisher’s exact test was used to test for the effect of specific clinical parameters, or treatments, on Ig positivity **(Table 1 and Figures 4 and 5**. Correlation of Ig levels with clinical parameters was tested by linear regression using Spearman correlation coefficient (R) (**Figure 5 and Supplemental Figure 1**). All statistical tests were carried out using established R scripts.

**Figure 2.**
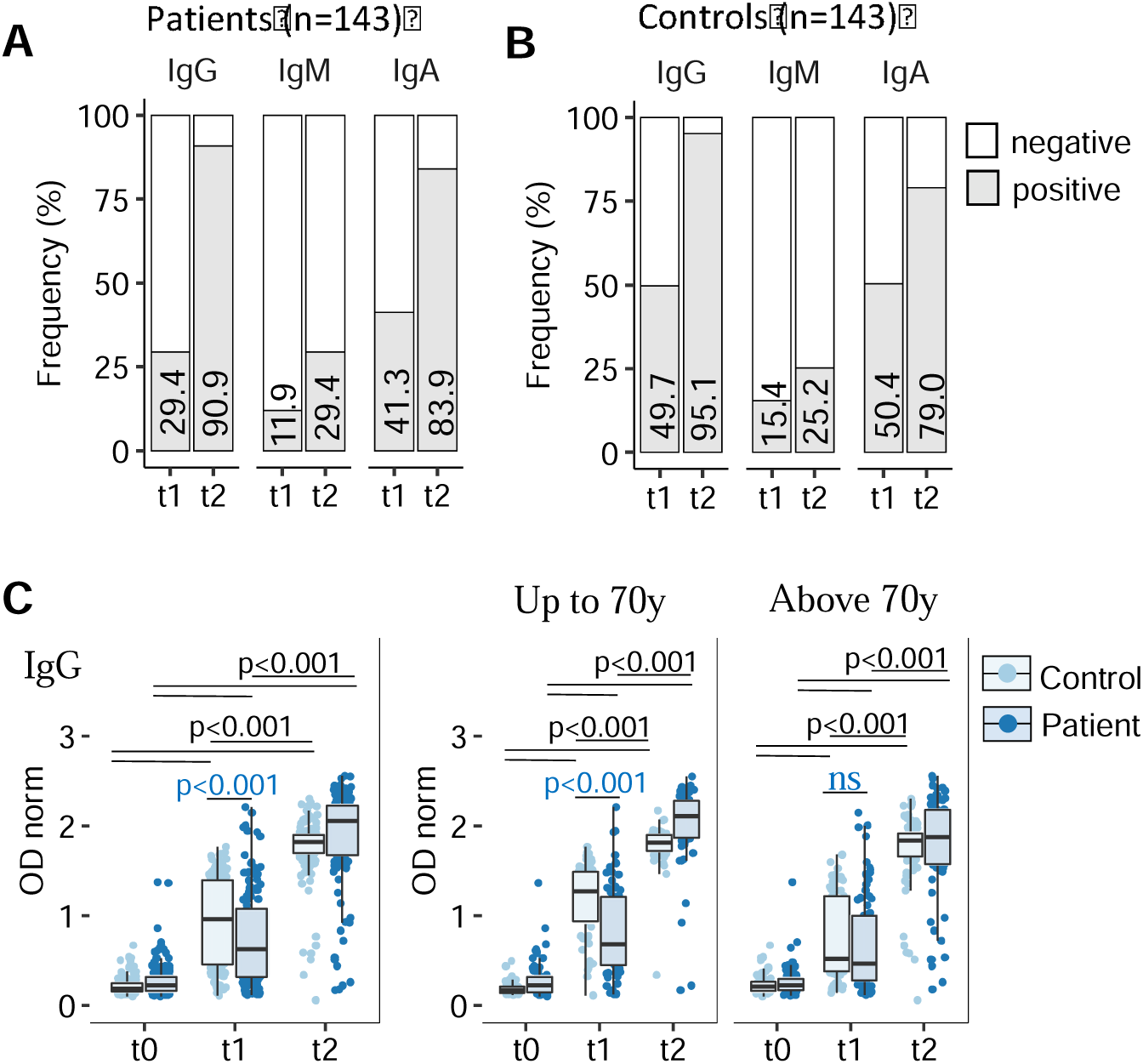
Heterogenous anti-SARS-CoV-2-spike responses to vaccination in hemodialyzed patients. Sera collected as described In Figure 1B (t0, t1 and t2) were analyzed for anti-full-length spike protein IgG, IgM and IgA antibodies (ELISA) in patients (n=143) and age-matched controls (n=143) A). Seroconversion defined by frequency of samples testing positive (grey bar) at t1 or t2 is represented for each antibody class in patients A) and controls B). ODnorm≥1 was used as cut-off for positivity. Seroconversion values are indicated inside each bar. C) Semi-quantitative analysis of anti full-length spike protein IgG in all patients (dark blue) and controls (light blue), or stratified by age up to 70, or above 70 years of age. Data points represent individual subjects and are overlaid with boxes representing interquartile range (IQR), whiskers representing 1.5 IQR tails, and median value. Increase of antibody levels across time was evaluated by Quade test for each antibody class (p-value <0.001), and significant Wilcoxon signed-rank test for pairwise comparison with or without age stratification, p-values indicated by horizontal black bars. Wilcoxon rank sum comparison of controls and patients at t1 are indicated with horizontal blue lines.

**Figure 3.**
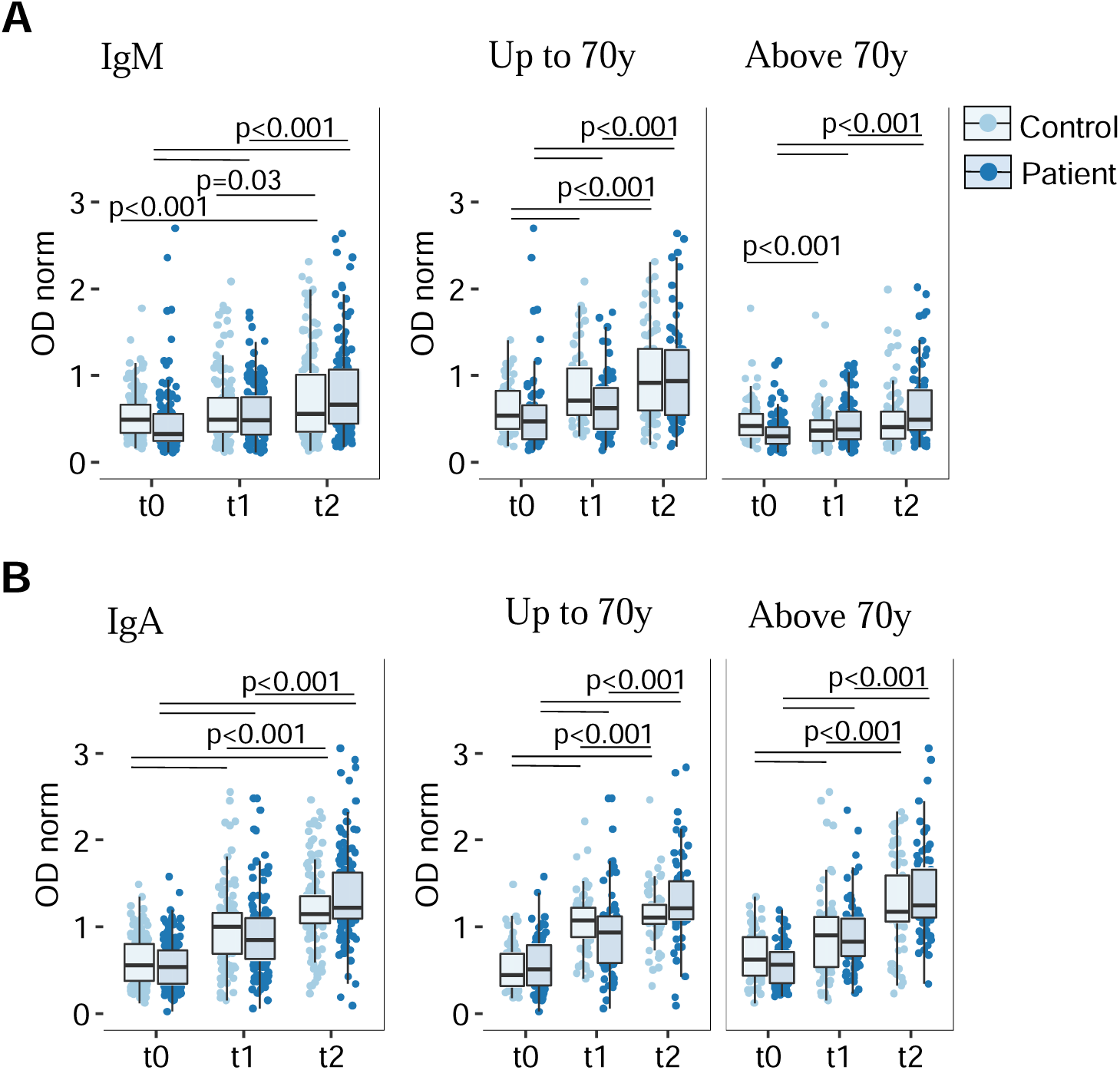
Anti-SARS-CoV-2-spike IgM and IgA responses to vaccination in hemodialyzed patients. Sera collected as described Figure 1B (t0, t1 and t2) were analyzed by ELISA for anti-full-length spike protein IgM (A) and IgA (B) in patients (n=143) and age-matched controls (n=143): in all patients (dark blue) and controls (light blue) (left), or stratified by age up to 70 (middle), or above 70 years of age (right). Data presentation and statistics as described in Figure 2C.

**Figure 4.**
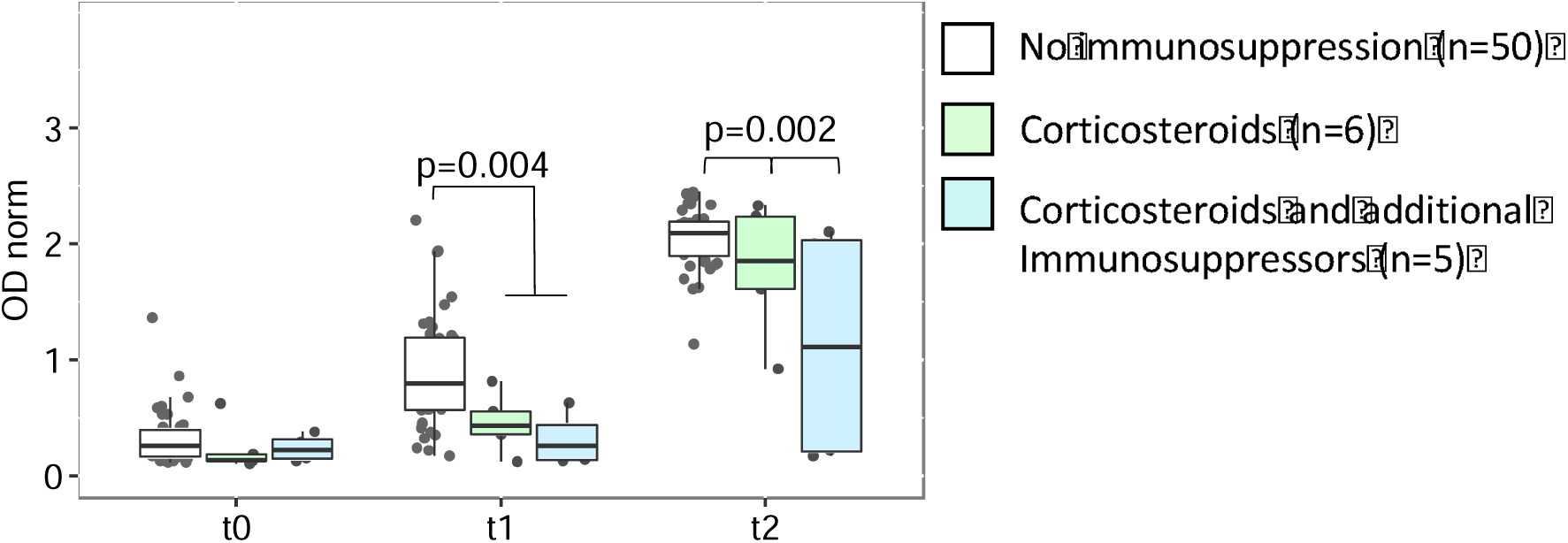
Anti-SARS-CoV-2- spike IgG reactivity induced by vaccination is lower in patients treated with immunosuppressors. Semi-quantitative analysis of IgG, IgM and IgA antibody levels stratified by immunosuppression treatment. The plot compares patients under corticosteroid treatment alone (green), patients treated with corticosteroids plus other immunosuppressors (blue) and 50 age-matched (40-69y) non-treated patients (in white). Data points represent individual subjects and are overlaid with boxes representing interquartile range (IQR), whiskers representing 1.5 IQR tails, and median value. Immunosuppression results in lower IgG levels at t1 and t2 (Wilcoxon rank sum exact test and Fisher’s exact test respectively, p-values are indicated).

**Figure 5.**
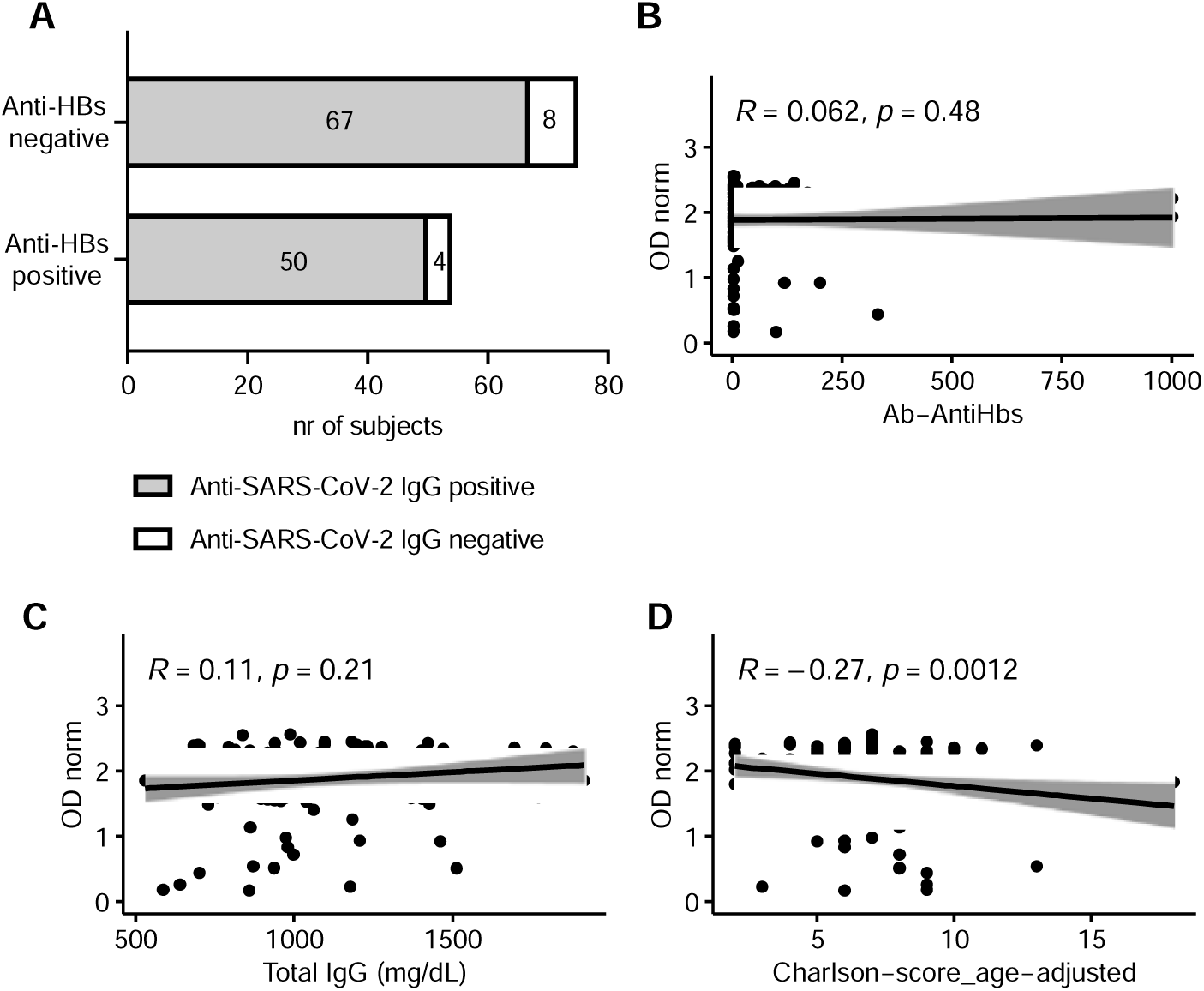
Analysis of anti-Spike IgG conditional to humoral response to HBs vaccine, serum IgG and co-morbidities in hemodialysis patients. A) Anti-spike positivity at t2 in 54 responders and 75 non-responders to previous hepatitis B vaccination (anti-HBs antibody cut-off > 10 mIU/mL). Fischer’s test p value=0.53; OR=1.49, 95%CI [0.48 to 4.65]. 14 Anti-Hbc reactive (previously infected with HBV) were excluded. Correlation analysis of anti-Spike IgG with anti-HBs levels in Ab-Anti Hbc not reactive individuals (n=129) (B) and total serum IgG (n=142) (C) as determined at t2. Correlation of anti-Spike IgG at t2 with age-adjusted Charlson comorbidity index (n= 142) (D). Shaded areas represent 95% Confidence Interval.

**Figure 6.**
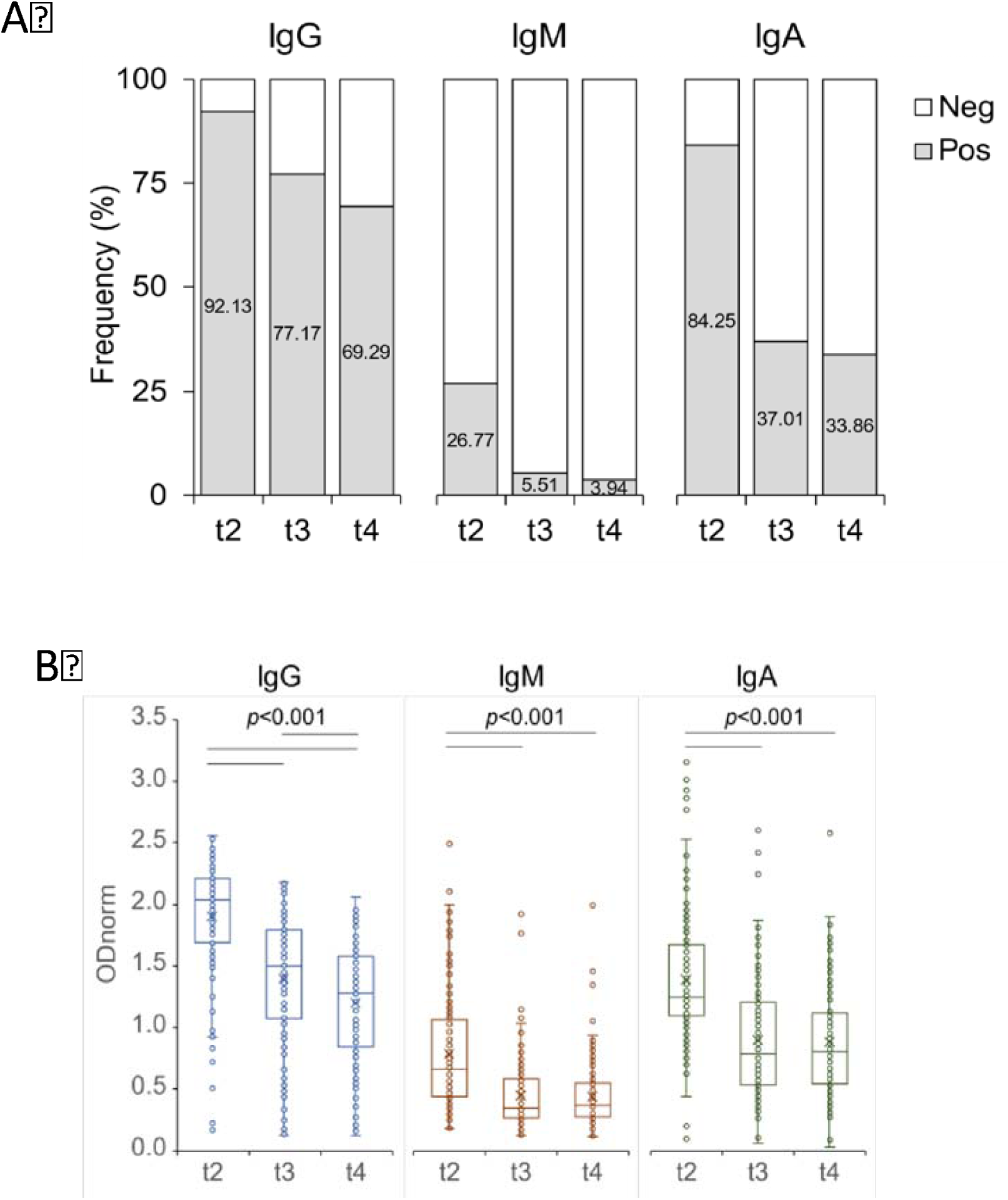
Decay of circulating antibodies in hemodialized patients up to 6 months after initiation of vaccination. Follow-up analysis of antibody responses for anti-full-length spike protein IgG, IgM and IgA antibodies (ELISA) in patients (n=127) collected at t2, t3 (140 days post-1^st^ vaccine dose) and t4 (180 days post-1^st^ vaccine dose). A) Seroconversion defined by frequency of samples testing positive (grey bar) is represented for each antibody class in patients. ODnorm≥1 was used as cut-off for positivity. Seroconversion values are indicated inside each bar. X-square statistics: t2 versus t3 for IgG (P=0.0009), IgM (p<0.0001), IgA (p<0.0001); t3 versus t4, IgG, IgM or IgA (P>0.1). B) Semi-quantitative analysis of anti full-length spike protein antibodies in patients. Data points represent individual subjects and are overlaid with boxes representing interquartile range (IQR), whiskers representing 1.5 IQR tails, and median value. Single comparisons of antibody levels across time were evaluated by Mann Whitney test for each antibody class. P-value of significant differences are indicated by horizontal black bars.

**Table 1:**
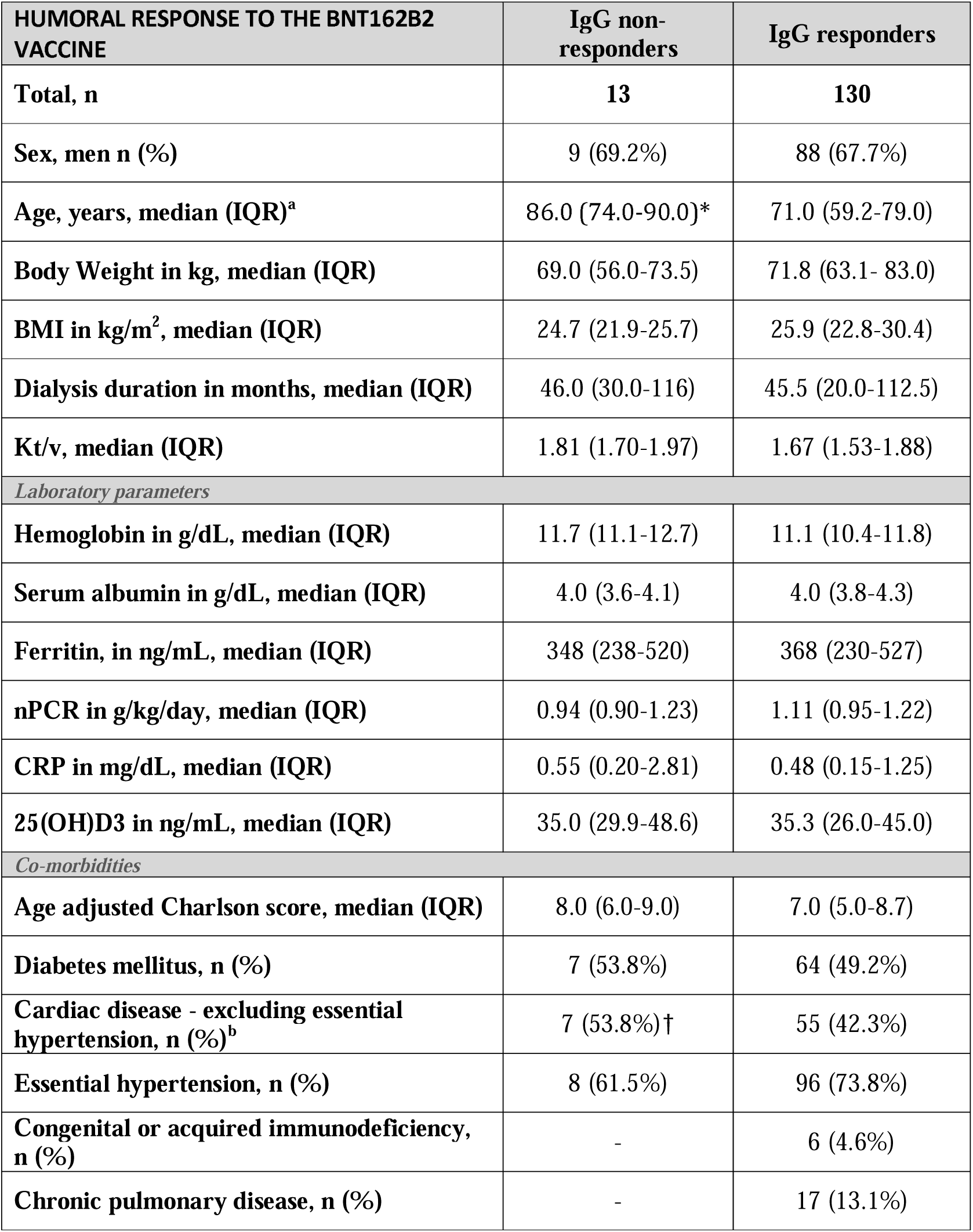
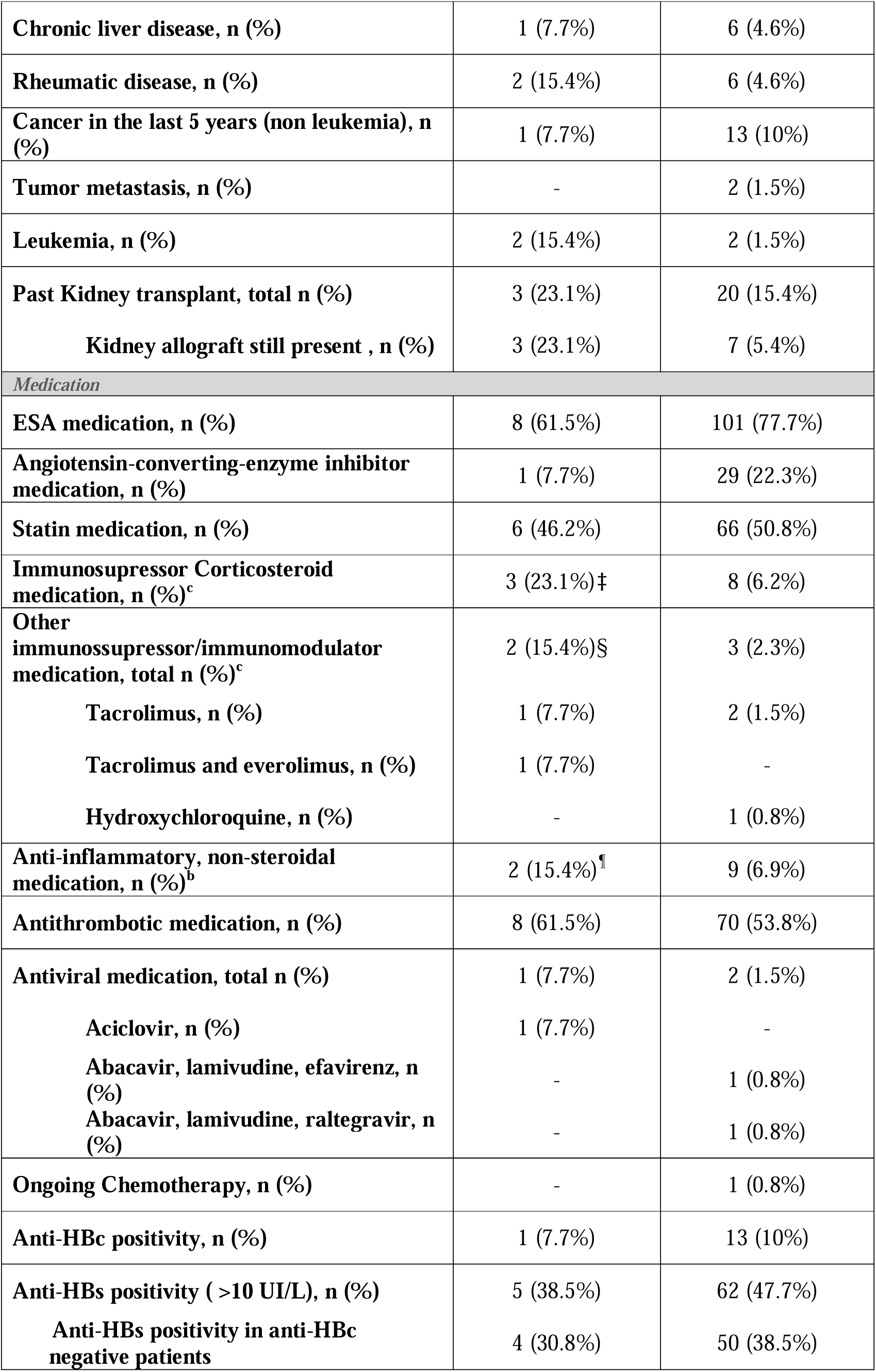
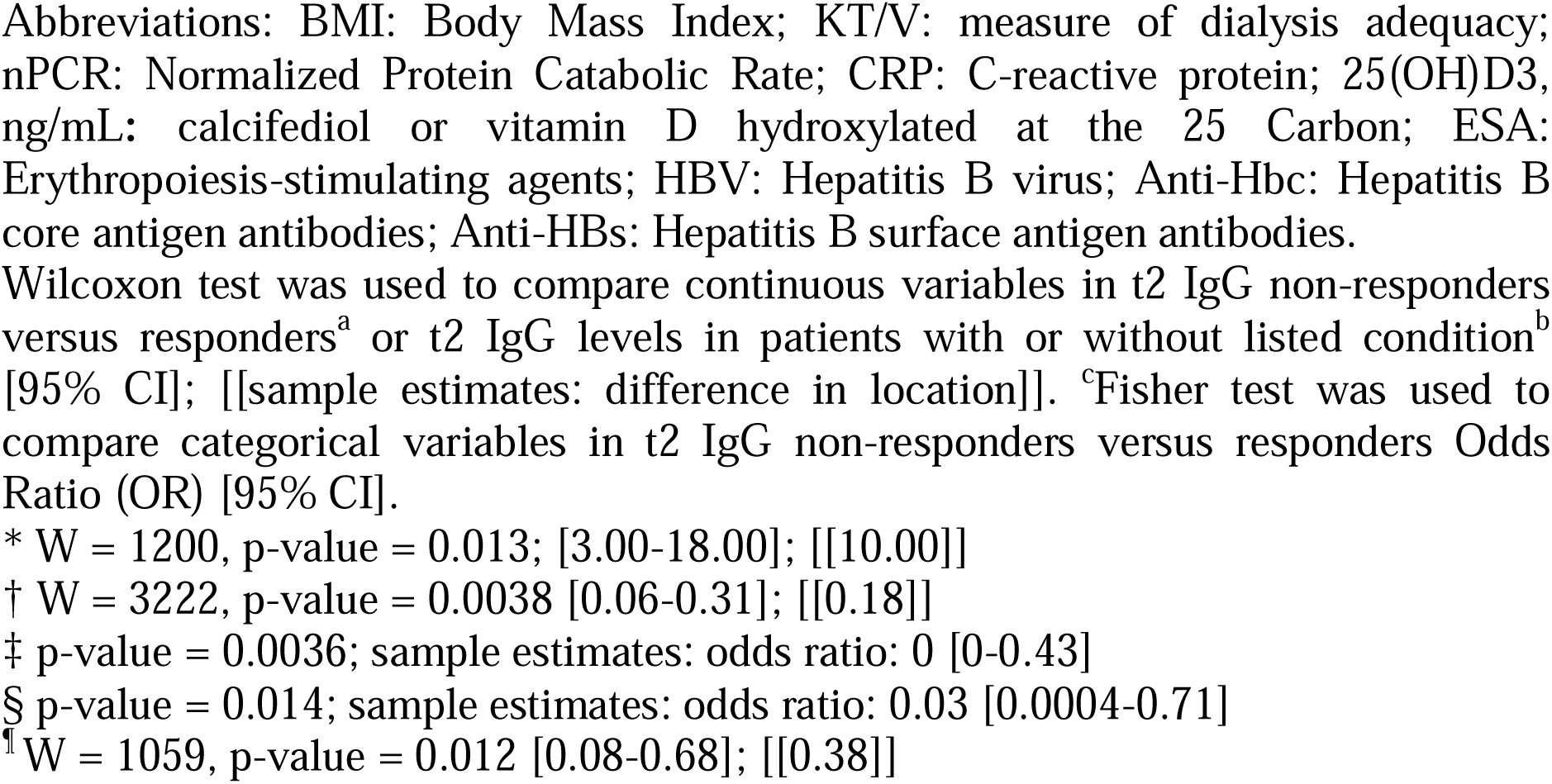
Clinical characterization of patients that did not generate anti-full-length Spike IgG antibodies (non-responders) in comparison with patients that did generate these IgG antibodies (responders) after two doses of Pfizer BNT162b2 vaccination (t2).

| HUMORAL RESPONSE TO THE BNT162B2 VACCINE | IgG non-responders | IgG responders |
| --- | --- | --- |
| **Total, n** | **13** | **130** |
| **Sex, men n (%)** | 9 (69.2%) | 88 (67.7%) |
| **Age, years, median (IQR)<sup>a</sup>** | 86.0 (74.0-90.0)* | 71.0 (59.2-79.0) |
| **Body Weight in kg, median (IQR)** | 69.0 (56.0-73.5) | 71.8 (63.1- 83.0) |
| **BMI in kg/m<sup>2</sup>, median (IQR)** | 24.7 (21.9-25.7) | 25.9 (22.8-30.4) |
| **Dialysis duration in months, median (IQR)** | 46.0 (30.0-116) | 45.5 (20.0-112.5) |
| **Kt/v, median (IQR)** | 1.81 (1.70-1.97) | 1.67 (1.53-1.88) |
| *Laboratory parameters* |  |  |
| **Hemoglobin in g/dL, median (IQR)** | 11.7 (11.1-12.7) | 11.1 (10.4-11.8) |
| **Serum albumin in g/dL, median (IQR)** | 4.0 (3.6-4.1) | 4.0 (3.8-4.3) |
| **Ferritin, in ng/mL, median (IQR)** | 348 (238-520) | 368 (230-527) |
| **nPCR in g/kg/day, median (IQR)** | 0.94 (0.90-1.23) | 1.11 (0.95-1.22) |
| **CRP in mg/dL, median (IQR)** | 0.55 (0.20-2.81) | 0.48 (0.15-1.25) |
| **25(OH)D3 in ng/mL, median (IQR)** | 35.0 (29.9-48.6) | 35.3 (26.0-45.0) |
| *Co-morbidities* |  |  |
| **Age adjusted Charlson score, median (IQR)** | 8.0 (6.0-9.0) | 7.0 (5.0-8.7) |
| **Diabetes mellitus, n (%)** | 7 (53.8%) | 64 (49.2%) |
| **Cardiac disease - excluding essential hypertension, n (%)<sup>b</sup>** | 7 (53.8%)† | 55 (42.3%) |
| **Essential hypertension, n (%)** | 8 (61.5%) | 96 (73.8%) |
| **Congenital or acquired immunodeficiency, n (%)** | - | 6 (4.6%) |
| **Chronic pulmonary disease, n (%)** | - | 17 (13.1%) |
| **Chronic liver disease, n (%)** | 1 (7.7%) | 6 (4.6%) |
| **Rheumatic disease, n (%)** | 2 (15.4%) | 6 (4.6%) |
| **Cancer in the last 5 years (non leukemia), n (%)** | 1 (7.7%) | 13 (10%) |
| **Tumor metastasis, n (%)** | - | 2 (1.5%) |
| **Leukemia, n (%)** | 2 (15.4%) | 2 (1.5%) |
| **Past Kidney transplant, total n (%)** | 3 (23.1%) | 20 (15.4%) |
| **Kidney allograft still present , n (%)** | 3 (23.1%) | 7 (5.4%) |
| *Medication* |  |  |
| **ESA medication, n (%)** | 8 (61.5%) | 101 (77.7%) |
| **Angiotensin-converting-enzyme inhibitor medication, n (%)** | 1 (7.7%) | 29 (22.3%) |
| **Statin medication, n (%)** | 6 (46.2%) | 66 (50.8%) |
| **Immunosuppressor Corticosteroid medication, n (%)<sup>c</sup>** | 3 (23.1%)<sup>‡</sup> | 8 (6.2%) |
| **Other immunosuppressor/immunomodulator medication, total n (%)<sup>c</sup>** | 2 (15.4%)<sup>§</sup> | 3 (2.3%) |
| **Tacrolimus, n (%)** | 1 (7.7%) | 2 (1.5%) |
| **Tacrolimus and everolimus, n (%)** | 1 (7.7%) | - |
| **Hydroxychloroquine, n (%)** | - | 1 (0.8%) |
| **Anti-inflammatory, non-steroidal medication, n (%)<sup>b</sup>** | 2 (15.4%)<sup>¶</sup> | 9 (6.9%) |
| **Antithrombotic medication, n (%)** | 8 (61.5%) | 70 (53.8%) |
| **Antiviral medication, total n (%)** | 1 (7.7%) | 2 (1.5%) |
| **Aciclovir, n (%)** | 1 (7.7%) | - |
| **Abacavir, lamivudine, efavirenz, n (%)** | - | 1 (0.8%) |
| **Abacavir, lamivudine, raltegravir, n (%)** | - | 1 (0.8%) |
| **Ongoing Chemotherapy, n (%)** | - | 1 (0.8%) |
| **Anti-HBc positivity, n (%)** | 1 (7.7%) | 13 (10%) |
| **Anti-HBs positivity ( >10 UI/L), n (%)** | 5 (38.5%) | 62 (47.7%) |
| **Anti-HBs positivity in anti-HBc negative patients** | 4 (30.8%) | 50 (38.5%) |

Abbreviations: BMI: Body Mass Index; KT/V: measure of dialysis adequacy; nPCR: Normalized Protein Catabolic Rate; CRP: C-reactive protein; 25(OH)D3, ng/mL: calcifediol or vitamin D hydroxylated at the 25 Carbon; ESA: Erythropoiesis-stimulating agents; HBV: Hepatitis B virus; Anti-Hbc: Hepatitis B core antigen antibodies; Anti-HBs: Hepatitis B surface antigen antibodies.
Wilcoxon test was used to compare continuous variables in t2 IgG non-responders versus responders<sup>a</sup> or t2 IgG levels in patients with or without listed condition<sup>b</sup> [95% CI]; [[sample estimates: difference in location]]. <sup>c</sup>Fisher test was used to compare categorical variables in t2 IgG non-responders versus responders Odds Ratio (OR) [95% CI].
\* W = 1200, p-value = 0.013; [3.00-18.00]; [[10.00]]
† W = 3222, p-value = 0.0038 [0.06-0.31]; [[0.18]]
‡ p-value = 0.0036; sample estimates: odds ratio: 0 [0-0.43]
§ p-value = 0.014; sample estimates: odds ratio: 0.03 [0.0004-0.71]
<sup>¶</sup> W = 1059, p-value = 0.012 [0.08-0.68]; [[0.38]]

## RESULTS

### COHORT CHARACTERIZATION

This longitudinal, prospective, cohort study enrolled 156 patients on hemodialysis, scheduled for BNT162b2 mRNA vaccination in January and February 2021; 143 participants adhered to the three collection times (**Figure 1A and B**). The median age was 72 years of age (y) (27-93) and females represented 32% of the cohort (**Figure 1C and D**). Eleven patients (8.8%) were under therapies potentially affecting immune responses (including corticosteroids, immunosuppressors and chemotherapy) (**Table 1)**. The control cohort included 143 age-matched individuals with median age of 73y (30-96) and 53,1% females (**Figure 1C and D**).

### ANTI-SPIKE ANTIBODY RESPONSES

Sera from hemodialyzed patients and controls were analyzed for specific anti-SARS-CoV-2-Spike antibodies (IgG, IgM and IgA) using ELISA calibrated with sera from COVID-19 patients, allowing discrimination of positive/negative antibody reactivity **(Figure 2A, B and Supplemental Table 1)**. This classification showed that 130/143 (90.9%; 95% CI 85.1-94.6) hemodialyzed patients and 136/143 (95,1%; 95%CI 90.2-97.6) controls developed anti-Spike IgG antibodies after the second vaccine dose (t2). After a single vaccine dose (t1), seroconversion was markedly lower in hemodialyzed patients with only 42/143 (29.4%; 95%CI 22.5-37.3) patients developing anti-Spike IgG antibodies when compared to 71/143 (49,7%; 95%CI 41.6-57.7) controls. Isotype class analysis of anti-spike antibodies also revealed marked progression in IgA seroconversion on hemodialyzed patients from t1 (41.3%; 95%CI, 33.5-49.5) to t2 (83.9%; 95%CI, 77.0-89.0), similar to the control cohort (50,4%, 95%CI 42.2-58.4 at t1 and 79%, 95%CI 71.6-84.9 at t2). In contrast, IgM antibodies showed low prevalence and modest increase along the vaccination schedule in both patients (11.9%; 95%CI 7.6-18.2 at t1 and 29.4%; 95%CI 22.5-37.3 at t2) and controls (15,4%, 95%CI 10.4-22.2 at t1 and 25,2%, 95%CI 18.8-32.9 at t2).

Semi-quantitative analysis of antibody levels using normalized OD values (ODnorm) showed significant increase of all three isotypes from t0 to t1 in both patients and controls, an effect of the first vaccine dose that was further enhanced by the second vaccine dose (t2) **(Figure 2C, Figure 3 and Supplemental Table 2)**. In particular, anti-spike IgG antibody levels in patients at t2 (median 2.05 and IQR [1.67-2.23]) were significantly higher when compared to t1 (0.63 [0.32-1.08]). Albeit to a lower extent, anti-spike IgA levels also increased from t1 (0.85 [0.63-1.10]) to t2 (1.22 [1.10-1.63]). Anti-spike IgM was only modestly increased from t1 (0.49 [0.32-0.75]) to t2 (0.66 [0.45-1.07]).

Comparison of anti-spike antibody levels in patients and controls revealed significant lower IgG levels in hemodialyzed patients after the first vaccine dose (0.63 [0.32-1.08], in patients, and 0.96 [0.46-1.39] in controls), an effect not observed for the other isotypes. At t2 the antibody levels are similar in patients and in controls for all the three isotypes **(Figure 2C and Figure 3)**.

To explore the effect of age on humoral response to vaccination we divided both cohorts into two age groups, below 70y and above 70y **(Figure 2C and Figure 3)**. We observed that elderly individuals present overall lower anti-spike IgG levels at t1, with equivalent responses in patients and controls (0.47 [0.28-1.00] in patients, and 0.52 [0.38-1.21] in controls, above 70). Remarkably, below 70y, the IgG response after the first vaccine dose (t1) was significantly lower in hemodialyzed patients (0.68 [0.45-1.21] when compared to controls (1.27 [0.93-1.49]), resembling the IgG levels observed in the elderly group. At t2, the IgG levels were similar in both age groups among patients and controls (**Supplemental Table 2**). An age effect was also observed in IgM at t1 and t2, although no difference was observed between the patient and control groups. In contrast, IgA antibody levels were not significantly affected by age (**Figure 3 and Supplemental Table 2**).

### ANTI-SPIKE IgG RESPONSE IN IMMUNOSUPPRESSED PATIENTS

Analysis of 9 patients under immunosuppression and 43 age-matched control patients (40-69 years) revealed that treatment significantly decreased anti-Spike IgG antibodies elicited by the first vaccine dose (median; IQR: immunosuppressed 0.36; IQR [0.14-0.63] controls 0.80; [0.6-1.21]; p-value=0.005). After the second dose (t2), 5 patients under corticosteroids therapy (1.85; [1.77-2.24]) and 4 patients under combined immunosuppression regimens (1.16; [0.20-2.24]) show lower anti-Spike IgG levels as compared to non-treated patients (2.12[1.95-2.28], p-value=0.008) **(Figure 4)**.

### ANTI-SPIKE IgG RESPONSE CORRELATION WITH HEPATITIS B VACCINATION, TOTAL SERUM IgG AND CO-MORBIDITIES

Clinical data was scrutinized to search for determinants of immuno-responsiveness to the BNT162b2 mRNA vaccination in hemodialyzed patients. Comparative analysis with response to hepatitis B vaccination was performed after exclusion of anti-HBc positive subjects, which indicates previous contact with the hepatitis B virus. Information on responsiveness to hepatitis B vaccination was available for 129 hepatitis B-naïve patients with only 54 (42%) patients maintaining anti-HBs positivity after vaccination (**Table 1**). In this sub-group 117/129 (91%) showed positivity to anti-Spike IgG at t2. Unresponsiveness to HBs immunization was not correlated with absence of anti-Spike IgG seroconversion after BNT162b2 mRNA vaccination (p value=0.53; OR=1.49, 95% CI [0.48-4.65]) **(Figure 5A)**. Similarly, anti-HBs antibodies levels did not correlate with anti-Spike IgG reactivity at t2 (r=0.062, p=0.48) **(Figure 5B)**.

Total IgG levels measured in 142 patients at t2, were not correlated with anti-Spike IgG levels (r=0.13, p-value=0.13) (**Figure 5C**). Likewise, total IgA and total IgM levels did not correlate with anti-Spike antibody responses (data not shown). Finally, we found weak but significant correlation (r=-0.27, p=0.001) between the age-adjusted Charlson comorbidity index and anti-Spike IgG levels at t2 (**Figure 5D**).

### CLINICAL CHARACTERIZATION OF IgG NON-RESPONDERS TO BNT162b2 mRNA VACCINE at t2

Reviewing clinical data of the thirteen subjects that remained anti-Spike IgG negative at t2 did not show significant over-representation of clinical conditions (**Table 1**).

Among non-responders, seven out of thirteen patients were older than 85 years of age. Among three non-responders less than 60 years old, two had functioning allografts under immunosuppression. Nevertheless, the majority of them did not present laboratory clues to inflammation, anemia, low serum albumin, 25-hydroxycholecalciferol levels or normalized protein catabolic rate (nPCR); clinical indicators including time in dialysis, KT/V and several inflammatory and nutritional parameters only reveal weak or non-significant effects on anti-Spike IgG levels (**Supplemental Figure 1**).

The presence of allograft and related use of immunosuppression was overrepresented in IgG non-responders (one kidney and one liver allograft recipients) (**Table 1**). Conversely, twelve additional kidney allograft recipients who underwent nephrectomy and stopped immunosuppression responded to the BNT162b2 mRNA vaccine.

### TIME COURSE OF ANTI-SPIKE RESPONSES FOLLOW-UP IN PATIENTS

To follow-up anti-spike antibody responses in patients on hemodialysis we further analyzed IgG, IgM and IgA antibodies in 127 patients (Figure 6). We found that IgG levels show a consistent decaying trend between t2 (42 days after first vaccine dose) and t3 (140 days) as well as between t3 and t4 (180 days). Nevertheless, the majority of patients (69.3%) still show IgG positivity 180 days after vaccine schedule initiation. On the other hand, IgM and IgA antibodies show an abrupt decay of antibody levels and sharp positivity drop by 140 days (t3) after vaccination initiation as compared to t2. These results indicate that after BNT162b2 mRNA vaccination patients on hemodialysis show IgG antibody responses that are more prevalent and more persistent as compared to other Ig classes, namely IgA. In addition, the results suggest that anti-spike IgG slow waning may lead to progressive decrease in positivity.

## DISCUSSION

Our results reveal that seroconversion following BNT162b2 initial dose is markedly lower in hemodialyzed patients when compared to reported antibody responses in the general population (8,10) and further indicate that 9% of these patients remain seronegative after the second vaccine dose. It is uncertain if other protective mechanisms are activated, for how long responders will be able to maintain adequate antibody titers and if additional doses are efficient in generating protection in non-responders. Therefore, these results support further investigation into the relationship between vaccination, serologic response and host protection.

Several studies unveiled an abnormal immune response both to viral infection and to vaccination in patients with CKD requiring renal replacement therapy (11–13). Blunted responses to influenza (14), pneumococcal (15) and hepatitis B vaccination (16) are paradigms of abnormal adaptative immunity in these patients. The effectiveness of hepatitis B vaccination protocols in conferring immune protection ranges between 51-69% for patients with CKD (17). This lack of response is in part due to uremic toxins and dialysis procedure that may lead to impaired macrophage function, dysregulated cytokine synthesis, lymphopenia and alterations in B-lymphocyte function (18). We found that anti-Spike seroconversion after BNT162b2 mRNA vaccination was not correlated with unresponsiveness to hepatitis B (HBs) immunization, with the mRNA vaccine to SARS-CoV-2 being more effective in eliciting humoral responses. Our results are in accordance to a previous publication (19) and indicate that the well-known high failure rate of HBs recombinant protein immunization in hemodialyzed patients did not provide an explanation for cases of unresponsiveness to the BNT162b2 mRNA vaccination.

Our results indicated that the second BNT162b2 RNA dose was critical to boost the humoral response in hemodialyzed patients, particularly the IgG antibodies. A sizable but significantly lower percentage of patients mounted an efficient serum IgA response. It will be interesting to evaluate if a similar level of secretory IgA is present, as anti-Spike IgA responses with neutralizing capacity were reported in natural SARS-CoV-2 infection (20). Conversely, IgM response was quite modest in this cohort. It is possible that different vaccine strategies may impact differently the isotypic response although their clinical significance remains unclear.

Our observations added to mounting evidence that age has a significant negative effect on antibody response to the BNT162b2 vaccine (5,6,8). Notably, IgG levels after one vaccine dose were remarkably lower in hemodialysis patients when compared to age-matched controls below 70 years. It is well known that hemodialysis patients present increased risk of cardiovascular disease when compared to age-matched controls, mimicking features observed in individuals several decades older (21). Furthermore, most hemodialysis patients in our cohort present comorbidities correlated with poor outcomes upon COVID-19 including advanced age, frailty, diabetes mellitus, hypertension, cardiovascular disease and cancer (22). Together, our analysis strengthen the notion that the dynamics of anti-IgG responses across the vaccination schedule in hemodialyzed patients resembles those of elderly cohorts (8). As expected, we observed a significant correlation of decreased humoral responses in the patients under immunosuppression at the time of vaccination and analysis. Our finding also support that patients on a waiting list for kidney transplantation have a better chance of developing an effective response to the vaccine while on hemodialysis than after transplantation (23,24). Although, our observations pertain a limited number of patients under immunosuppressive therapies they support that antibody responses should be followed-up in those patients.

Many concerns remain regarding the efficacy of the immune response elicited by vaccination against SARS-Cov-2 in hemodialyzed patients (25). We have further explored our hemodialyzed cohort to evaluate whether non-responders could have specific features worth addressing in future vaccination strategies. Total IgG levels were not associated with anti-Spike responses. The detected association signals of heart disease, rheumatic disease, usage of antithrombotic agents, use of non-steroidal anti-inflammatory drugs and cancer (including leukemia) in unresponsiveness to vaccine warrants confirmation in cohorts with higher sample size.

Overall, our results show that a small fraction of hemodialyzed patients do not reach positivity for anti-spike IgG after two vaccine doses and that age and immunosuppressive treatments are associated to decreased levels of anti-spike IgG in response to BNT162b2 RNA vaccine. These results are in line with other studies reporting that less than 100% hemodialysis patients have detectable antibodies (5,10,26). However, in some studies, previous exposition to SARS-Cov-2 was not excluded, as anti-N antibodies were not measured prior to vaccination (5) which could have inflated responsiveness estimates.

Within 7 months following the second dose of vaccination, only one patient from our cohort of patients on maintenance hemodialysis became infected with SARS-CoV2. This patient had been one of the 13 individuals on hemodialysis that did not present an antibody response to vaccine and was tested due to contact with a COVID-19 positive case. Although asymptomatic for COVID-19, this patient died from massive gastrointestinal hemorrhage. Larger cohorts of patients on hemodialysis will be necessary to evaluate the effectiveness of the vaccine on preventing death, morbidities and interpatient viral transmission.

In addition, we found significant antibody decay by 180 days after the beginning of vaccination that was less marked in the IgG class. Natural immunity acquired by contact of the virus may be long lasting, associated with persistence of antibodies providing partial protection against reinfection ((27,28). Whether vaccination is also associated with long term protection in patients on maintenance hemodialysis is uncertain at this point. Our observations warrant further studies to investigate if antibody waning correlates with increased risk of infection and symptomatic COVID-19 in patients on hemodialysis.

Cellular immunity and other potential protective mechanisms were not investigated in this study and may also be relevant for vaccine effectiveness (24).

To the best of our knowledge, this is the first report of such a dramatic fall in antibody titers in patients on hemodialysis, with potential implications on the duration of protection against SARS-CoV2 and raising the possible need of a booster dose, as suggested by other authors (29).

## Data Availability

The datasets generated during and/or analysed during the current study are available from the corresponding author on reasonable request

## Acknowledgments

João Frazão, MD, PhD for scientific approval and critical review of the manuscript; Ana Paula Agudo, MD, Gabriela Teixeira, MD, João Gomes, MD, Jorge Araújo, MD, Rui Costa, MD for data collection; Carla Brás for secretarial assistance; nursing staff of DaVita Obidos for sample collection; Affidea laboratories for sample management.

## Author’s Contributions

A.W., J.D. and C.P-G. conceived and designed the study and drafted the article. M-L.B, L.G., I.G. and N.D., collected and analyzed the data and drafted the article. R.A., P.B., A.B., V.M., P.M., O.A., L.K., P.C., E.N., M.P., A.F., TN and JV collected the data and carried out the assays. All authors revised and approved the final version of the manuscript.

