## Supplemental Material for "Longitudinal analysis of antibody responses to the Pfizer BNT162b2 vaccine in Patients Undergoing Maintenance Hemodialysis" (from another version of this preprint: version 1, 2021-07-22)

Table of contents:

**Supplemental Table 1** - Anti-Spike IgG, IgM and IgA positivity for data presented in Figure 2

**Supplemental Table 2** - Median and Inter quartile range [IQR] for semi-quantitative IgG, IgM and IgA data presented in Figure 2

**Supplemental Figure 1** - Correlation analysis of anti-Spike IgG at t2 with relevant pathology indicators.

**Supplemental Table 1.** Anti-Spike IgG, IgM and IgA positivity for data presented in Figure 2

| | | | | Positivity; n (%; 95% CI) | |
|---|---|---|---|---|---|
| | **Isotype** | **Age** | **n** | **t1** | **t2** |
| Patients | IgG | All | 143 | 42 (29.4; 22.5-37.3) | 130 (90.9; 85.1-94.6) |
| | | 27-70 | 66 | 22 (33.3; 23.2-45.3) | 63 (95.5; 87.5-98.4) |
| | | 71-93 | 77 | 20 (26.0; 17.5-36.7) | 67 (87.0; 77.7-92.8) |
| | IgM | 27-93 | 143 | 17 (11.9; 7.6-18.2) | 42 (29.4; 22.5-37.3) |
| | IgA | 27-93 | 143 | 59 (41.3; 33.5-49.5) | 120 (83.9;77.0-89.0) |
| Controls | IgG | All | 143 | 71 (49.7; 41.6-57.7) | 136 (95.1; 90.2-97.6) |
| | | 30-70 | 66 | 48 (72.7; 61.0-82.0) | 65 (98.5; 91.9-99.7) |
| | | 71-96 | 77 | 23 (29.9; 20.8-40.8) | 71 (92.2; 84.0-96.4) |
| | IgM | 30-96 | 143 | 22 (15.4; 10.4-22.2) | 36 (25.2;18.8-32.9) |
| | IgA | 30-96 | 143 | 72 (50.4; 42.2-58.4) | 113 (79.0;71.6-84.9) |

**Supplemental Table 2.** Median and Inter quartile range [IQR] for semi-quantitative IgG, IgM and IgA data presented in Figure 2

| | | | | | Median [IQR] | |
|---|---|---|---|---|---|---|
| | **Isotype** | **Age** | **n** | **t0** | **t1** | **t2** |
| Patients | IgG | All | 143 | 0.22 [0.16- 0.32] | 0.63 [0.32- 1.08] | 2.05 [1.67- 2.23] |
| | | 27-70 | 66 | 0.22 [0.15- 0.32] | 0.68 [0.45- 1.21] | 2.11 [1.87- 2.28] |
| | | 71-93 | 77 | 0.22 [0.17- 0.30] | 0.47 [0.28- 1.00] | 1.87 [1.57- 2.18] |
| | IgM | 27-93 | 143 | 0.33 [0.24- 0.56] | 0.49 [0.32- 0.75] | 0.66 [0.45- 1.07] |
| | IgA | 27-93 | 143 | 0.53 [0.35- 0.73] | 0.85 [0.63- 1.10] | 1.22 [1.10- 1.63] |
| Controls | IgG | All | 143 | 0.19 [0.15- 0.24] | 0.96 [0.46- 1.39] | 1.82 [1.70- 1.90] |
| | | 30-70 | 66 | 0.17 [0.14- 0.21] | 1.27 [0.93- 1.49] | 1.81 [1.72- 1.89] |
| | | 71-96 | 77 | 0.21 [0.16- 0.26] | 0.52 [0.38- 1.21] | 1.83 [1.66- 1.92] |
| | IgM | 30-96 | 143 | 0.49 [0.34- 0.66] | 0.49 [0.35- 0.75] | 0.56 [0.35- 1.01] |
| | IgA | 30-96 | 143 | 0.56 [0.38- 0.80] | 1.00 [0.69- 1.16] | 1.15 [1.04- 1.35] |

**Supplementary Figure 1**

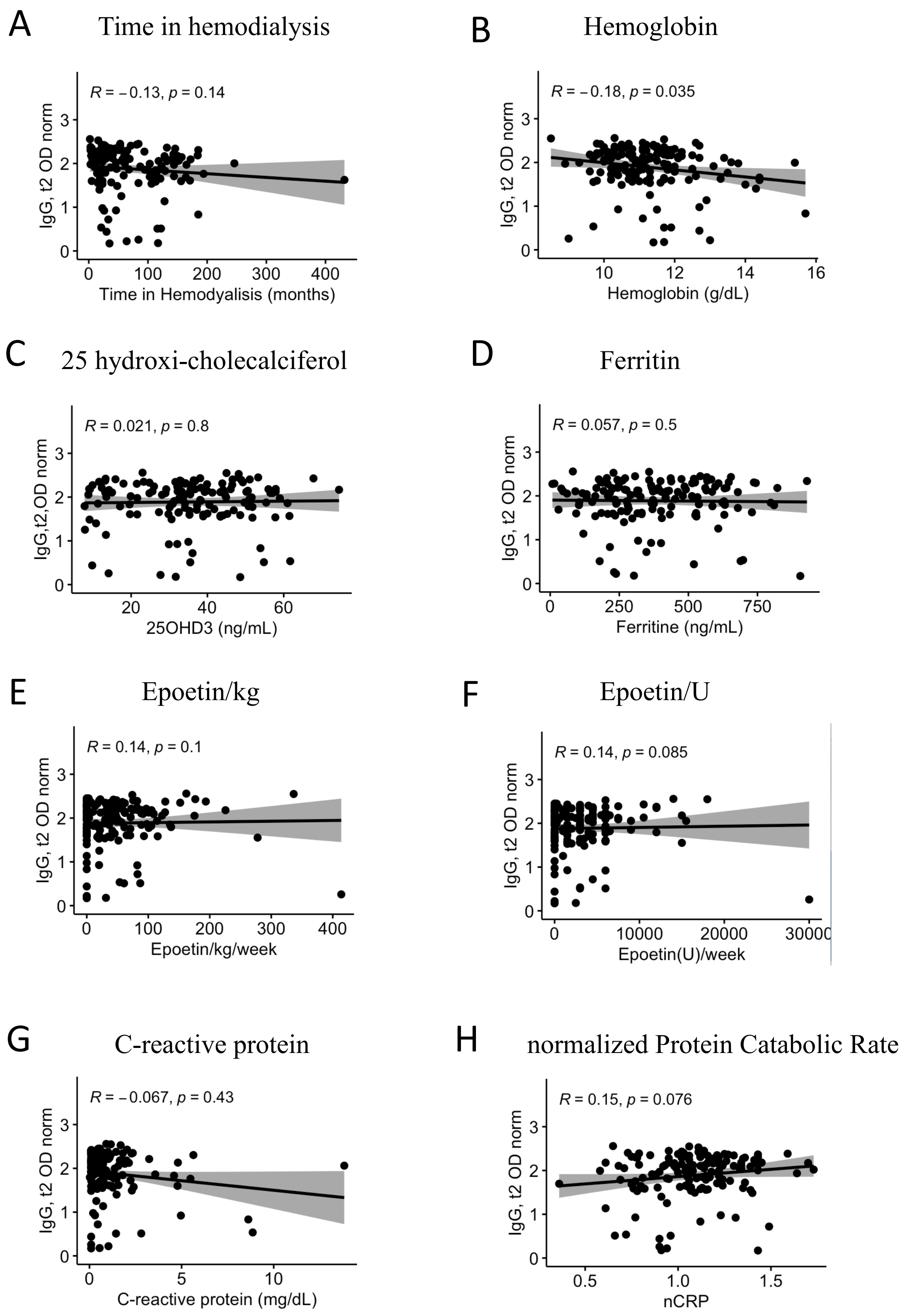

**Supplementary Figure 1:** Correlation analysis of anti-Spike IgG at t2 with relevant pathology indicators. Correlation with time in hemodialysis (A); with Hemoglobin levels (B); with 25 hydroxi-cholecalciferol (C); with Ferritin (D); with Epoetin/Kg (E) and Epoetin/U per week (F); with C-reactive protein (G) and normalized Protein Catabolic Rate (nPCR) (H).
